# Mitral Annular Disjunction in Out-of-Hospital Cardiac Arrest Patients – a Retrospective Cardiac MRI Study

**DOI:** 10.1101/2023.06.05.23290557

**Authors:** Felix Troger, Gert Klug, Paulina Poskaite, Christina Tiller, Ivan Lechner, Martin Reindl, Magdalena Holzknecht, Priscilla Fink, Eva-Maria Brunnauer, Elke R. Gizewski, Bernhard Metzler, Sebastian Reinstadler, Agnes Mayr

**Author notes:** Address for Correspondence: Agnes Mayr, MD University Clinic of Radiology Medical University of Innsbruck Anichstrasse 35, A-6020 Innsbruck, Austria.

## Abstract

**Background:** Mitral annular disjunction (MAD), defined as defective attachment of the mitral annulus to the ventricular myocardium, has recently been linked to malignant arrhythmias. However, its role and prognostic significance in patients requiring cardio-pulmonary resuscitation (CPR) remains unknown. This retrospective analysis aimed to describe prevalence and significance of MAD by cardiac magnetic resonance (CMR) imaging, in out-of-hospital cardiac arrest (OHCA) patients.

**Methods:** Eighty-six patients with OHCA and a CMR scan 5 days after CPR (interquartile range (IQR): 49 days before – 9 days after) were consecutively enrolled. MAD was defined as disjunction-extent ≥1mm in CMR long-axis cine-images. Medical records were screened for laboratory parameters, comorbidities and prior arrhythmias.

**Results:** In 34 patients (40%), no underlying cause for OHCA was found during hospitalization despite profound diagnostics. Unknown-cause OHCA patients showed a higher prevalence of MAD compared to definite-cause patients (56% vs. 10%, p<0.001) and had a MAD-extent of 6.3mm (IQR: 4.4-10.3); moreover, these patients were significantly younger (43 years vs. 61 years, p<0.001), more often female (74% vs. 21%, p<0.001) and had fewer comorbidities (hypertension, hypercholesterolemia, coronary artery disease, all p<0.005). By logistic regression analysis, presence of MAD remained significantly associated with OHCA of unknown cause (odds ratio: 8.49, 95% confidence interval: 2.37-30.41, p=0.001) after adjustment for age, presence of hypertension and hypercholesterolemia.

**Conclusions:** MAD is rather common in OHCA patients without definitive aetiology undergoing CMR. Presence of MAD remains independently associated to OHCA without identifiable trigger. Further research is needed to understand the exact role of MAD in OHCA patients.

**Clinical Perspectives:**

- This study showed that MAD is apparently a common finding in cardiac arrest patients without underlying trigger and was associated with it independently of age, concomitant hypertension and hypercholesterolemia.
- In clinical routine, MAD should be considered as potential arrhythmogenic substrate especially in those cardiac arrest patients, in which eventually no clear etiology can be found.
- However, future studies need to further explore the role of MAD in these patients and investigate the true arrhythmogenic potential of this anatomical variant.

## Introduction

Mitral annular disjunction (MAD) represents the defective anchoring of the mitral valve annulus into the ventricular myocardium (1). This anatomical variant has long been regarded as rather common, but clinically irrelevant secondary finding to mitral valve prolapse (MVP) (2). However, its status as a distinct disease entity acting as a possible substrate for ventricular arrhythmias has been increasingly substantiated within the past few years (3, 4). Recent studies suggested that its formerly assumed prevalence has been clearly underestimated (5); additionally, the term MAD was uncoupled from its status as a negligible auxiliary finding of MVP, as it was shown that MAD could be detected even without concomitant prolapse (3). To date, the research interest in MAD is continuously growing (2). However, data about its clinical relevance and postulated association to ventricular arrhythmias are scarce (6-8). Furthermore, data about its prevalence and significance in out-of-hospital cardiac arrest (OHCA) patients are completely lacking. Nevertheless, OHCA represents a leading cause of mortality worldwide (9), with an estimated 20% being of unknown or unobtainable cause (10). Although most studies tend to use echocardiography to screen and evaluate MAD, assessment by cardiac magnetic resonance (CMR) imaging seems more appropriate, especially in MAD of minor extent (11).

Accordingly, the aims of this retrospective study were as follows: a) to determine the prevalence of MAD in a population of OHCA patients; b) to assess its prevalence in OHCA patients in whom no definite cause of cardiac arrest (CA) was finally definable; and, c) to classify the role of MAD in this latter patient group.

## Methods

### Study population

The study population included all OHCA patients treated at the local university hospital from June 2007 to April 2021, where an adequate CMR scan was available. Patient records were screened for comorbidities and risk factors, positive family history for coronary artery disease (CAD) or CA and laboratory parameters as well as further diagnostic measures, including electrocardiography (ECG) and cardiac computed tomography (CT). Moreover, these records were checked for additional rhythmological events before, during or after hospital stay. CAD was defined as any coronary atherosclerotic disease detected in the respective modality (i.e. CT or cardiac catheterization). A flowchart of in- and excluded patients is shown in figure 2. This study was approved by the local Ethics Committee and conforms to the Declaration of Helsinki.

**Figure 1.**
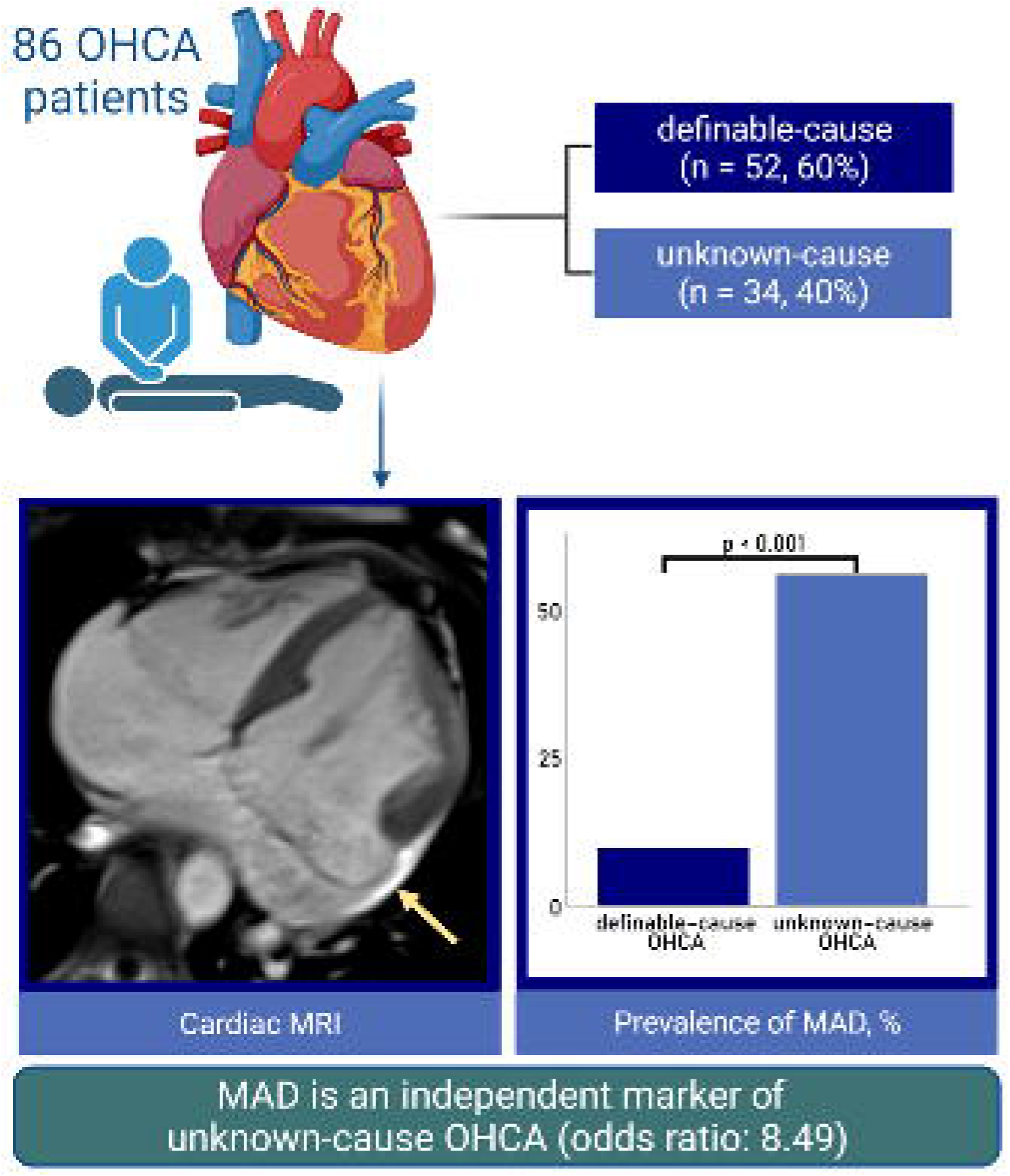
Study synopsis - mitral annular disjunction occurs frequently in unknown-cause out-of-hospital cardiac arrest and represents an independent marker after adjustment for age, hypertension and hypercholesterolemia. (Illustration created with biorender.com). *MAD: mitral annular disjunction, OHCA: out-of-hospital cardiac arrest*.

**Figure 2.**
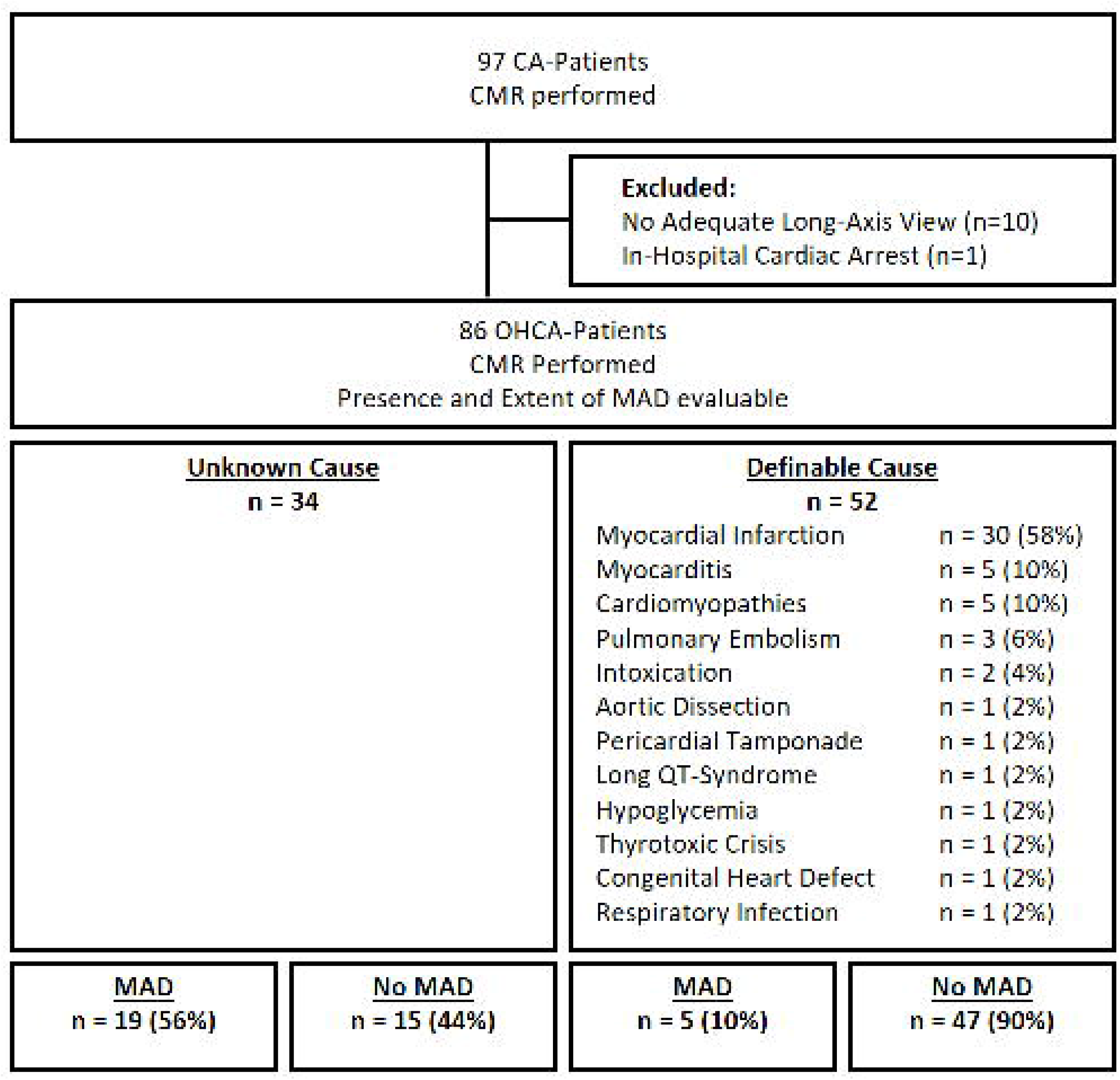
Flowchart of in- and exclusion, displaying the definable causes of CPR. *CA: cardiac arrest, CMR: cardiac magnetic resonance imaging, CPR: cardio-pulmonary resuscitation, MAD: mitral annular disjunction, OHCA: out-of-hospital cardiac arrest*.

### Cardiovascular Magnetic Resonance Imaging

All CMR scans were performed on a 1.5 Tesla clinical MR scanner (MAGNETOM Avanto or Avantofit; Siemens Healthineers AG, Erlangen, Germany). The standard CMR-protocol included high-resolution cine-images in long- and short axis view covering the left ventricle (LV), using a balanced steady state free precession sequence with retrospective ECG-gating (slice thickness: 8mm, interslice gap: 2mm, echo time: 1.19ms, repetition time: 2.83ms, 22 lines per segments, median temporal resolution: 39.9 ms (interquartile range (IQR): 38.4-46.1), frame rate: 25 frames per second, flip angle: 70°, field of view: 380×310mm, matrix: 320×260, voxel size: 2.6×1.8×8.0mm³, parallel imaging mode: GRAPPA (generalized autocalibrating partial parallel acquisition) with acceleration factor 2). ECG-triggered, phase-sensitive inversion recovery sequences were used to obtain late gadolinium enhancement (LGE) images 15-20 minutes after application of a 0.2 mmol/kg body mass gadolinium-contrast bolus. Standard software (Circle Cardiovascular Imaging, Calgary, Canada) was used for post-processing-analyses with semi-automatic detection of LV and right ventricular endo- and epicardial borders. Papillary muscles were excluded from myocardial mass (MM) and included in the LV volume. End-diastolic volume (EDV) and end-systolic volume (ESV) were then divided by the body surface area (BSA) [m²] to obtain indexed values (EDVi and ESVi). To calculate BSA, the Du Bois-formula was used (12).

MAD was defined as the presence of detachment ≥1mm between the mitral annulus and the ventricular myocardium, affecting the area under the posterior valve leaflet (3). Extent of MAD was measured longitudinally as distance from atrial valve leaflet junction to the top of the LV myocardium, at end-systole, in long-axis cine-images (figure 4). Only patients with a CMR of sufficient quality to decide whether MAD is present or not were included in this study, in order to avoid false-positive diagnoses of MAD (so called ‘pseudo-MAD’ that is feigned by juxtaposition of the posterior leaflet (13)). To determine the particular affected mitral segments, suitable short-axis slices were used.

MVP was defined as superior displacement ≥2mm of any part of the mitral leaflet beyond the mitral annulus (3, 14). Systolic curling motion was defined as unusual systolic motion of the posterior mitral ring on the adjacent myocardium (15), as illustrated in figure 3. MAD-presence and –extent as well as presence of MVP were conducted in full by two independent observers blinded to clinical data, each with several years of experience in CMR diagnosis (AM, 13 years, EuroCMR level II certified and FT, 3 years).

**Figure 3.**
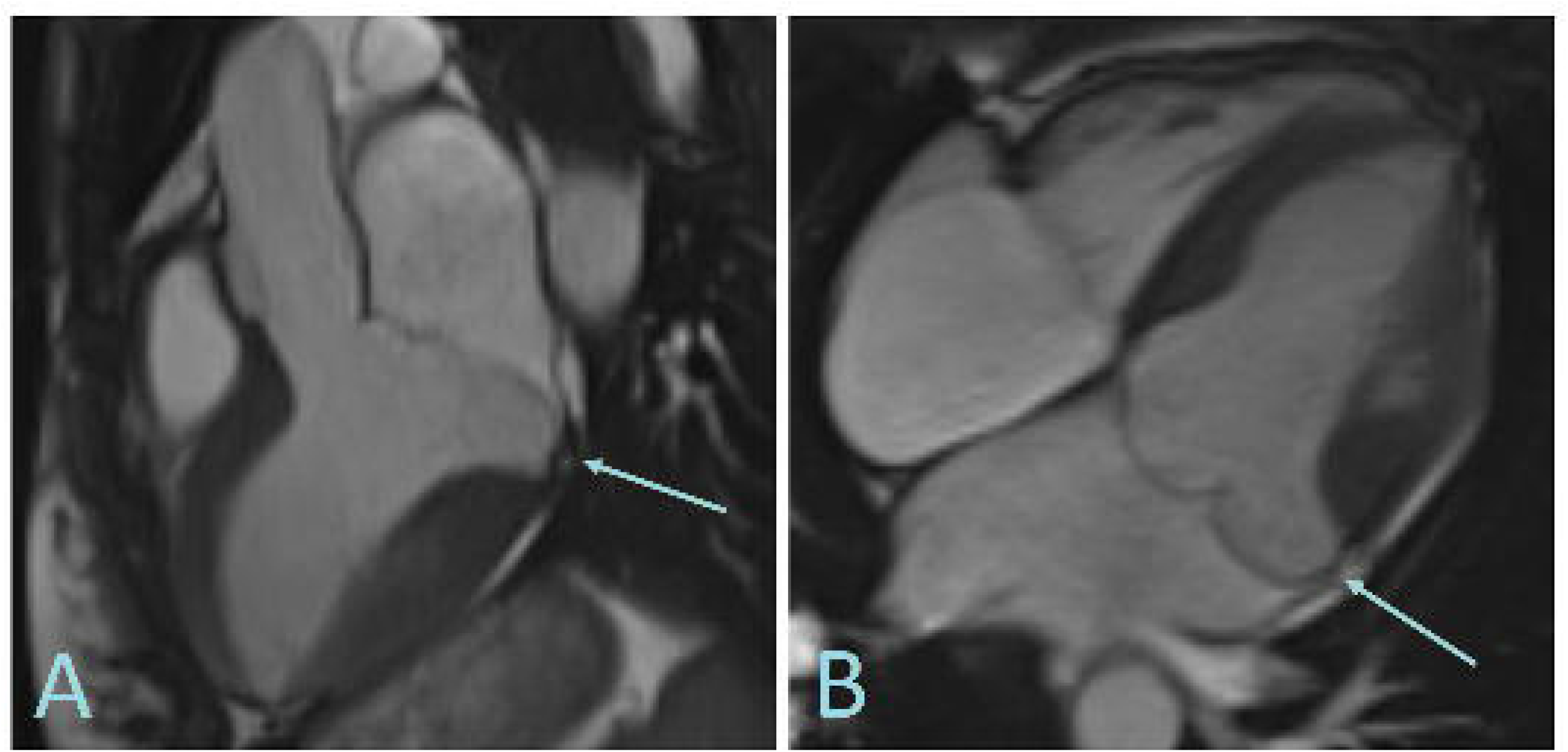
Three- (A) and four-chamber (B) view of a female patient in her 20s with out-of-hospital cardiac arrest of unknown cause, showing a distinct mitral annular disjunction with a systolic curling (arrow) of the P2- and P1-segment of the mitral valve with a maximum extent of 14.2mm.

**Figure 4.**
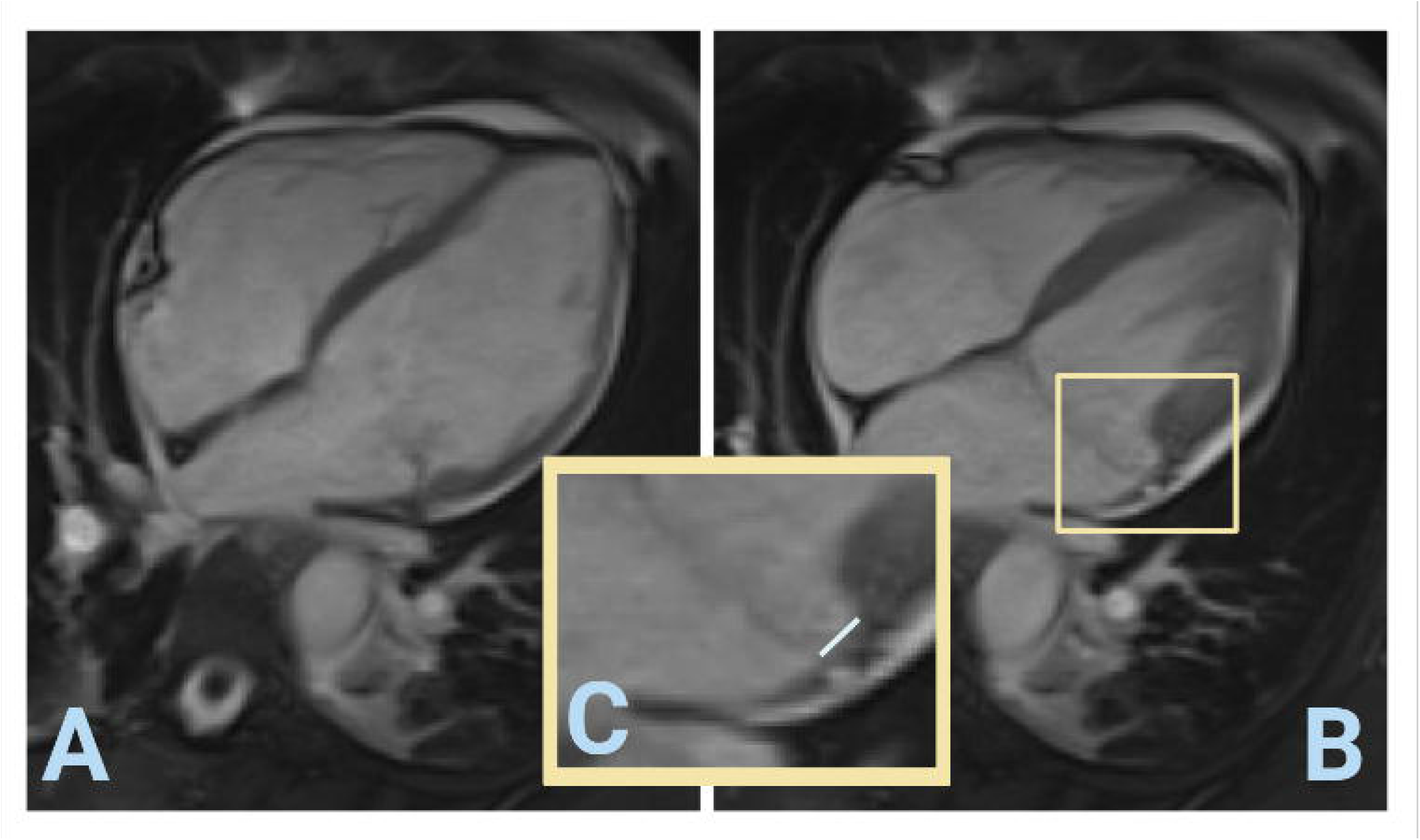
The detection of mitral annular disjunction and the measurement of its extent depends very much on the correct phase: this patient showed a marked disjunction of the P1-segment in end-systole (B), which is not visible at end-diastole (A). Panel (C) shows the positioning of the measuring tool as was used in this current study.

To quantify LGE, ‘hyper-enhancement’ was defined as ≥5 standard deviations above the signal intensity of remote myocardium in the opposite segment of the left ventricle. LGE was then measured on consecutive short-axis slices, infarct size is presented as a percentage of LV myocardial mass.

### Statistical Analysis

SPSS Statistics 26.0 (IBM, Armonk, NY, USA) was used for statistical analyses. All results for continuous variables are expressed as medians with corresponding IQR, categorical variables as absolute numbers and percentages. Differences in continuous and categorical variables between two groups were tested by Mann-Whitney U-test and chi-square test, respectively. A p-value <0.05 was considered as statistically significant. Logistic regression analysis was performed to evaluate independent markers for OHCA of unknown cause as well as independent markers for MAD; variables with a p-value <0.10 in univariable analysis and within these the variables of clinical relevance were included in our multivariable models.

## Results

### Baseline patient characteristics

For the current retrospective analysis, 86 OHCA patients were included. All patients underwent cardio-pulmonary resuscitation (CPR), with a median age of 56 years (IQR: 41-67). CMR was performed 5 days after resuscitation (IQR: 49 days before – 9 days after). At hospital discharge, no definite reason for CA was found after excluding coronary/cardiac, infectious, thromboembolic, genetic/congenital or metabolic conditions as well as intoxications, in 34 patients (40%). These patients are referred to as ‘unknown-cause OHCA’. Patient inclusion criteria as well as the particular causes for CA are listed in figure 2. Baseline characteristics are shown in table 1.

**Table 1.**
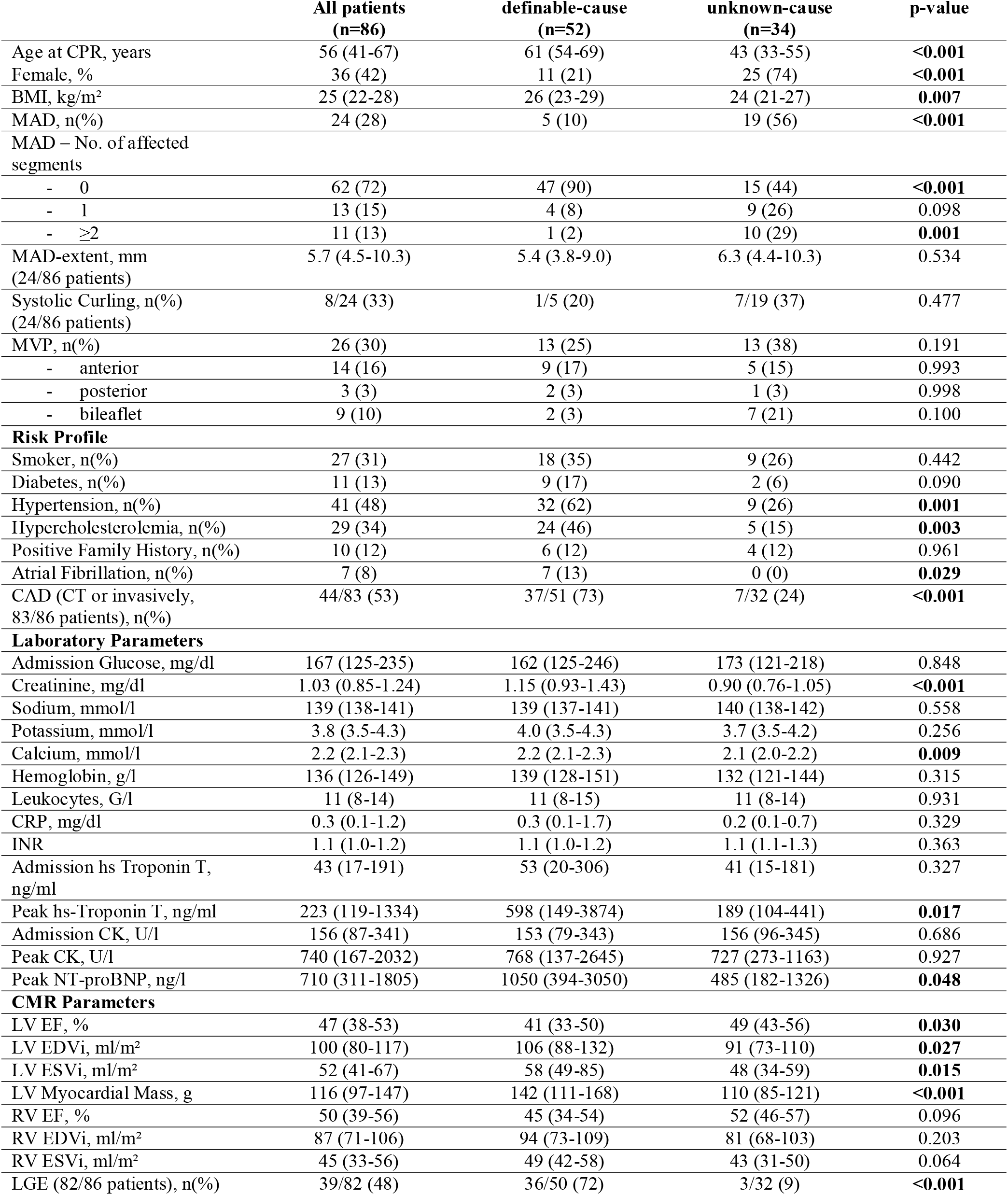
Baseline characteristics. *BMI: body mass index, CAD: coronary artery disease, CK: creatine kinase, CPR: cardio-pulmonary resuscitation, CRP: C-reactive protein, CT: computed tomography, EDVi: indexed end-diastolic volume, EF: ejection fraction, ESVi: indexed end-systolic volume, hs: high-sensitive, INR: international normalized ratio, LGE: late gadolinium enhancement, LV: left ventricular, MAD: mitral annular disjunction, MVP: mitral valve prolapse, NT-proBNP: N-terminal pro-B-type natriuretic peptide, RV: right ventricular*.

### Mitral Annular Disjunction

Overall, MAD was present in 28% of OHCA patients (n=24), with a median MAD-extent of 5.7mm (IQR: 4.5-10.3), ranging from 2.8 to 14.3mm. Patients with MAD were significantly younger (40 years [IQR: 32-52] vs. 61 years [IQR: 50-70], p<0.001) and more often female (75% vs. 29%, p<0.001). Moreover, MAD-patients had a lower BMI (23kg/m² [IQR: 20-26] vs. 26kg/m² [IQR: 23-29], p=0.006) and a lower prevalence of diabetes, hypertension and hypercholesterolemia (all p<0.03). Within MAD-patients, 8 showed systolic curling motion (33%). MVP was present in 26 patients (30%; with MAD: n=15 [63%] vs. without MAD: n=11 [18%], p<0.001). A detailed comparison of patients with and without MAD is shown in table 2.

**Table 2.** Comparison of patients with and without MAD. *BMI: body mass index, CAD: coronary artery disease, CK: creatine kinase, CPR: cardio-pulmonary resuscitation, CRP: C-reactive protein, CT: computed tomography, EDVi: indexed end-diastolic volume, EF: ejection fraction, ESVi: indexed end-systolic volume, hs: high-sensitive, INR: international normalized ratio, LGE: late gadolinium enhancement, LV: left ventricular, MAD: mitral annular disjunction, MVP: mitral valve prolapse, NT-proBNP: N-terminal pro-B-type natriuretic peptide, RV: right ventricular*.

|  | **MAD**<br>**(n=24)** | **No MAD**<br>**(n=62)** | **p-value** |
| --- | --- | --- | --- |
| Age at CPR, years | 40 (32-52) | 61 (50-70) | **<0.001** |
| Female, % | 18 (75) | 18 (29) | **<0.001** |
| BMI, kg/m<sup>2</sup> | 23 (20-26) | 26 (23-29) | **0.006** |
| MVP, n(%) | 15 (63) | 11 (18) | **<0.001** |
| - anterior | 7 (29) | 7 (11) | 0.261 |
| - posterior | 1 (4) | 2 (3) | 0.998 |
| - bileaflet | 7 (29) | 2 (3) | **0.007** |
| **Risk Profile** |  |  |  |
| Smoker, n(%) | 6 (25) | 21 (34) | 0.375 |
| Diabetes, n(%) | 0 (0) | 11 (18) | **0.024** |
| Hypertension, n(%) | 5 (21) | 36 (58) | **0.001** |
| Hypercholesterolemia | 3 (13) | 26 (42) | **0.007** |
| Positive Family History, n(%) | 2 (8) | 8 (13) | 0.523 |
| Atrial Fibrillation, n(%) | 0 (0) | 7 (11) | 0.078 |
| CAD (CT or invasively,<br>83/86 patients), n(%) | 4/23 (17) | 40/60 (67) | **<0.001** |
| **Laboratory Parameters** |  |  |  |
| Admission Glucose, mg/dl | 166 (120-202) | 167 (125-246) | 0.368 |
| Creatinine, mg/dl | 0.84 (0.72-1.01) | 1.13 (0.92-1.41) | **<0.001** |
| Sodium, mmol/l | 140 (138-141) | 139 (137-142) | 0.315 |
| Potassium, mmol/l | 3.7 (3.5-4.0) | 3.9 (3.5-4.3) | 0.240 |
| Calcium, mmol/l | 2.1 (2.0-2.2) | 2.2 (2.1-2.3) | **0.021** |
| Hemoglobin, g/l | 131 (117-139) | 140 (128-151) | **0.032** |
| Leukocytes, G/l | 12 (7-15) | 11 (8-14) | 0.836 |
| CRP, mg/dl | 0.1 (0.1-1.2) | 0.3 (0.1-1.2) | 0.238 |
| INR | 1.1 (1.0-1.3) | 1.1 (1.0-1.2) | 0.829 |
| Admission hs Troponin T,<br>ng/ml | 70 (14-187) | 37 (19-216) | 0.777 |
| Peak hs-Troponin T, ng/ml | 200 (126-681) | 267 (104-3175) | 0.399 |
| Admission CK, U/l | 187 (103-521) | 151 (79-333) | 0.164 |
| Peak CK, U/l | 921 (361-2067) | 504 (143-2032) | 0.182 |
| Peak NT-proBNP, ng/l | 402 (131-907) | 1064 (431-3034) | **0.003** |
| **CMR parameters** |  |  |  |
| LV EF, % | 47 (43-56) | 45 (33-52) | 0.111 |
| LV EDVi, ml/m<sup>2</sup> | 94 (80-111) | 103 (79-126) | 0.376 |
| LV ESVi, ml/m<sup>2</sup> | 49 (40-58) | 56 (41-77) | 0.158 |
| LV Myocardial Mass, g | 102 (85-118) | 125 (110-160) | **0.006** |
| RV EF, % | 54 (46-58) | 48 (34-54) | **0.043** |
| RV EDVi, ml/m<sup>2</sup> | 85 (71-104) | 89 (71-109) | 0.685 |
| RV ESVi, ml/m<sup>2</sup> | 43 (32-49) | 46 (33-58) | 0.129 |
| LGE (82/86 patients), n(%) | 5/24 (21) | 34/58 (59) | **0.002** |

### Unknown-Cause Out-of-Hospital Cardiac Arrest

Patients without definite substrate for CA were significantly younger (43 years [IQR: 33-55] vs. 61 years [IQR: 54-69], p<0.001) and more often female (n=25 [74%] vs. n=11 [21%], p<0.001). Nineteen OHCA patients without definite cause for CA had MAD (56%) with a median MAD-extent of 6.3mm (IQR: 4.4-10.3); of these, 10 patients (53%) with MAD had two or three mitral valve segments affected. Unknown-cause OHCA patients had a lower body mass index (BMI, 24kg/m² [IQR: 21-27] vs. 26kg/m² [IQR: 23-29], p=0.007]) and a lower prevalence of arterial hypertension (26% vs. 62%, p=0.001) and hypercholesterolemia (15% vs. 46%, p=0.003). Before hospitalization, atrial fibrillation occurred only in patients with a definite cause for CA (n=7, 13%).

A total of 83 OHCA patients (97%) were evaluated for the presence of any CAD, either by coronary angiography (performed in 63 patients (76%) on the day of CPR [IQR: 0-6 days after]) or by coronary computed tomography angiography (CTA) (in 52 patients (60%) performed on the day of CPR [IQR: 2 days before - 1 day after]). Combined, these two modalities resulted in an overall CAD-prevalence of 53%. CAD was significantly more common in patients with a definable cause for OHCA (73% vs. 24%, p<0.001).

MAD was shown to be significantly associated with unknown-cause OHCA univariably (odds ratio (OR): 11.91, 95% confidence interval (CI): 3.79-37.37, p<0.001) and to be an independent marker of unknown-cause OHCA after adjustment for age, hypertension and hypercholesterolemia (OR: 8.49, 95%CI: 2.37-30.41, p=0.001) by logistic regression analysis. Results of uni- and multivariable analysis are listed in table 3.

**Table 3.**
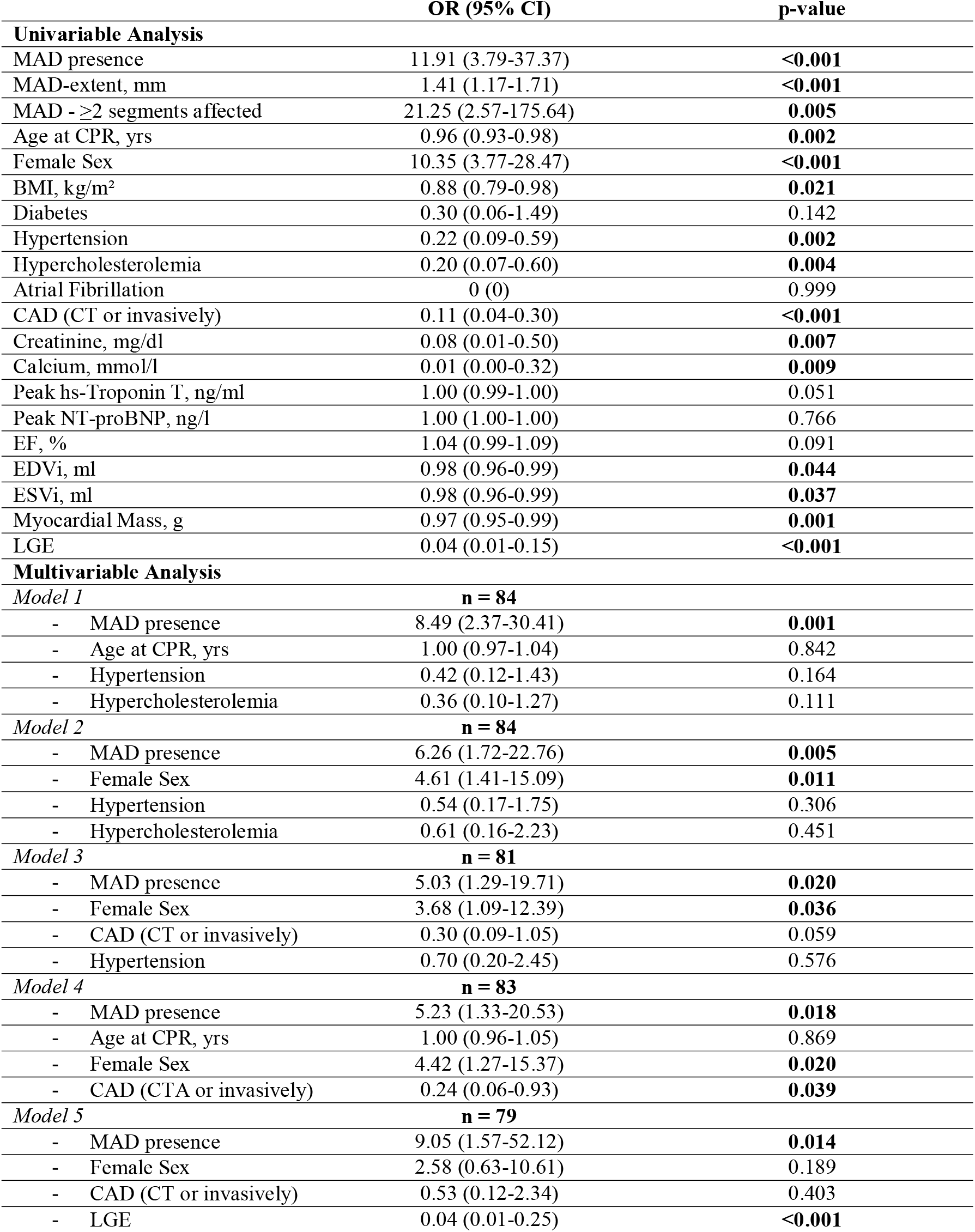
Uni- and multivariable analyses *BMI: body mass index, CAD: coronary artery disease, CPR: cardio-pulmonary resuscitation, CT: computed tomography, EDVi: indexed end-diastolic volume, EF: ejection fraction, ESVi: indexed end-systolic volume, hs: high-sensitive, LGE: late gadolinium enhancement, MAD: mitral annular disjunction, NT-proBNP: N-terminal pro-B-type natriuretic peptide.*

### CMR measurements

In unknown-cause OHCA patients (n= 34, 40%), CMR was performed 6 days after CPR (IQR: 6-8). In 6 of these, CMR was performed before CPR, with specific indications including evaluation of ventricular extrasystoles (n=2), tachyarrhythmia (n=2) or suspected (but eventually not confirmed) myocarditis (n=2). In the remaining 28 patients, CMR was performed in the course of diagnostic workup of CA.

In CMR, LV ejection fraction (EF) differed significantly between unknown-cause OHCA patients and those with a definite cause (49% [IQR: 43-56] vs. 41% [IQR: 33-50], p=0.030), as did EDVi, ESVi and MM (all p<0.03). Dichotomized at a median LV-EF of 49%, patients with a lower LV-EF had their scan 4.8 days after CPR, while the patient group with an LV-EF above the median had their scan 7.4 days after CPR (p=0.013). Furthermore, LV-EF correlated significantly with days between CPR and CMR (spearman’s rho: 0.574, p<0.001). Contrast agent was administered in 82 patients (95%). LGE was found in 39 patients (48%, 30 ischemic vs. 9 non-ischemic pattern), with unknown-cause OHCA patients presenting significantly less common with LGE (9% vs. 72%, p<0.001). An ischemic LGE pattern was found in 6% of unknown-cause patients (n=2/32, in both cases small-focal areas) and in 56% of definite-cause patients (n=28/50). Overall, MAD-patients showed LGE significantly less often (n=5/24, 21% vs. n=34/58, 59%, p=0.002) compared to patients without MAD presence. No MAD-patient showed papillary muscle enhancement. A detailed list of CMR measurements is shown in table 1 and 2, respectively.

Within unknown-cause OHCA patients, only 3/32 (9%) patients showed LGE at CMR, with two patients each presenting with a single small-focal ischemic LGE (one in the right coronary artery area (RCA) affecting 0.7% of LV-MM and one in the left anterior descending artery (LAD) affecting 2.1% of LV-MM), both likely due to thromboembolic events and both remained without clinical signs or abnormalities on coronary angiography. The one patient in this group showing a non-ischemic pattern had LGE of the basal posterolateral wall without affecting the papillary muscles (affecting 3.9% of LV-MM).

In the definite-cause OHCA group, 28/50 patients (56%) had ischemic LGE (culprit lesions: LAD – n=13 [46%]; RCA – n=8 [29%]; circumflex artery (CX) – n=2 [7%]; multiple – n=5 [18%]; median infarct size 11% of LV-MM, IQR: 7-21) and 8/50 patients had non-ischemic LGE (1.7% of LV-MM, IQR: 1.1-1.9) of the posterolateral wall without affecting any papillary muscle. Papillary muscle enhancement was found in 13% of patients (n=11, all of them with ischemic LGE; 1 unknown-cause [3%] vs. 10 definite-cause [20%], p=0.172) and affected the posterolateral papillary muscle in 10 patients (91%, 3 with both papillary muscles affected, 7 with isolated posterolateral papillary muscle enhancement).All patients with papillary enhancement had ischemic LGE, none of the MAD-patients had papillary muscle enhancement.

### Rhythmological Features

A detailed list of rhythmological features is shown in table 4. At the index event, the initial rhythm (recorded in 81 patients, 94%) showed no difference between OHCA patients with unknown cause and with definable cause (p=0.155) or between patients with and without MAD (p=0.051), with MAD-patients presenting exclusively with ventricular fibrillation (VF). Post-CPR-ECG on the day of index CA was available in 79 patients (92%) and differed significantly between unknown-cause OHCA patients and those with a definable cause concerning repolarization disorders (p=0.020), primarily concerning ST-elevation (12/47, 26% vs. 1/32, 3%), with the other entities encompassing unspecific repolarization disorders. There was no significant difference regarding rhythm (p=0.568), electrical heart axis (p=0.349), P-wave morphology (0.211), bundle branch blocks (p=0.337), pathological Q-waves (p=0.843), signs of hypertrophy (p=0.387) and specific time intervals (PQ, QRS, QT/QTc, all p>0.2).

**Table 4.**
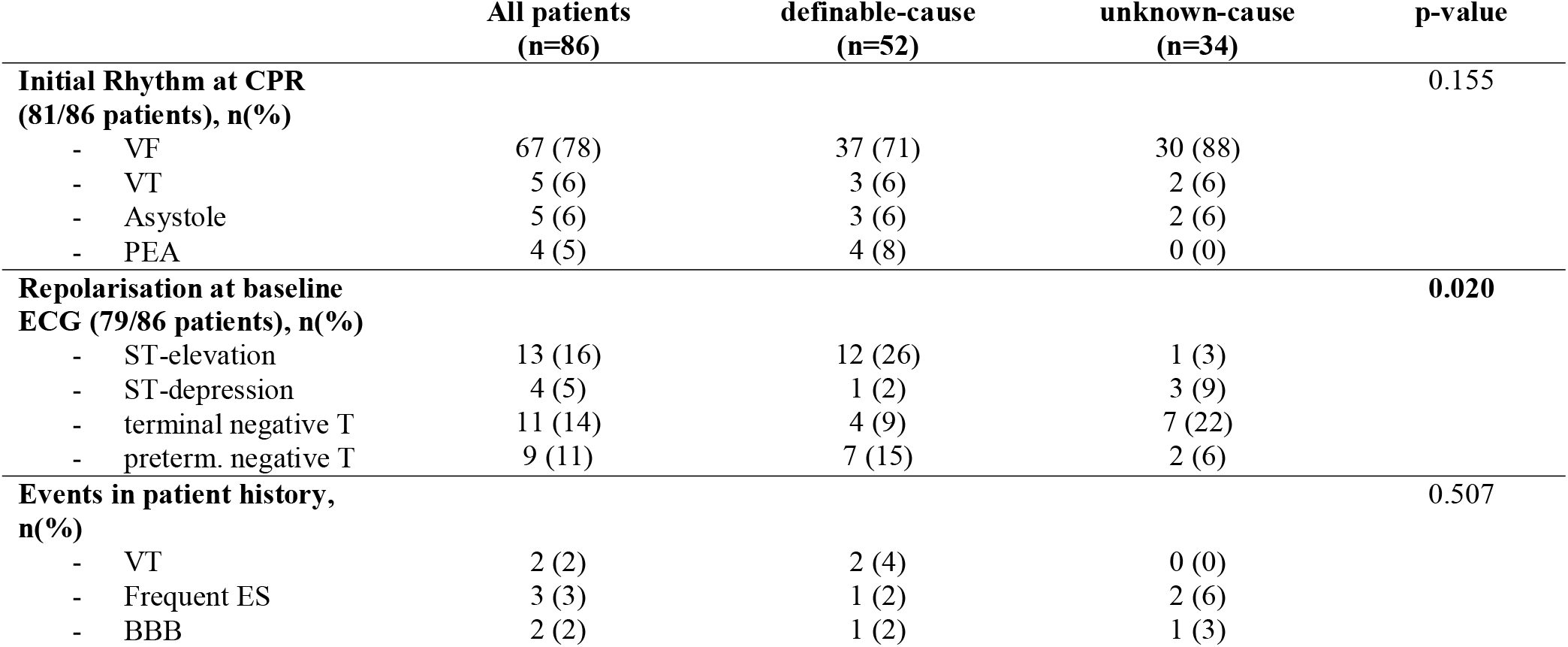
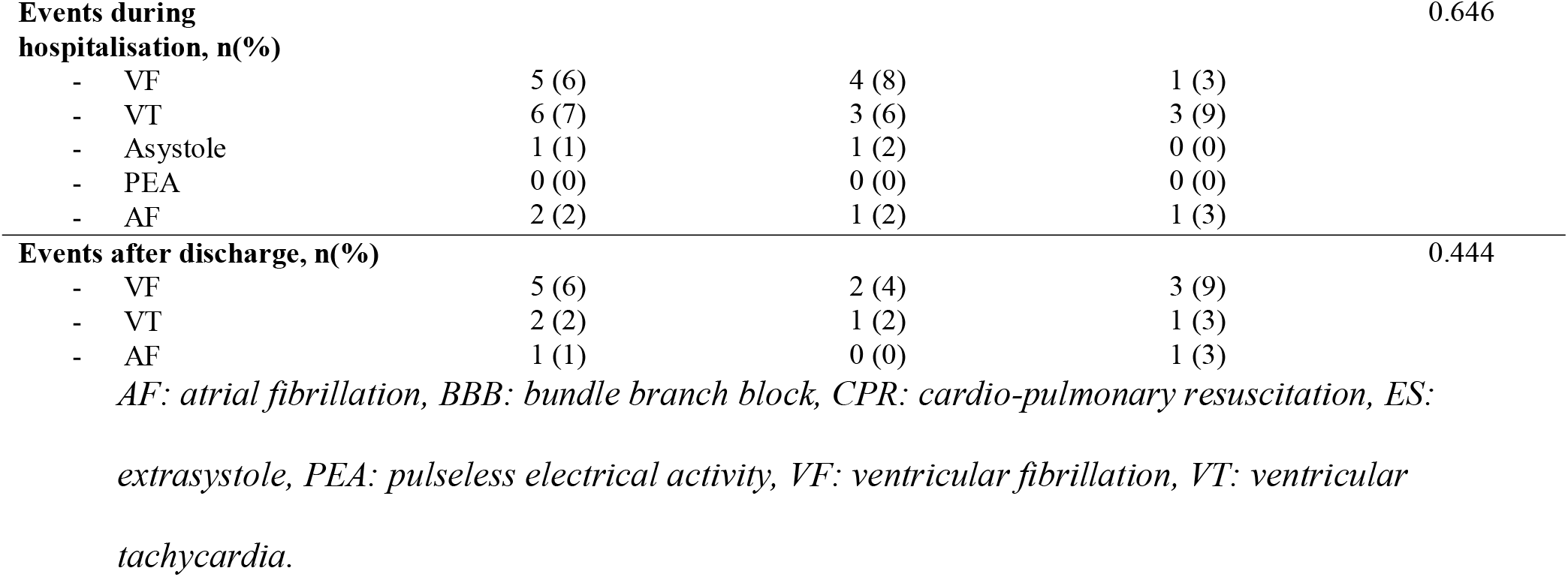
Rhythmological features.

|  | **All patients**<br>**(n=86)** | **definable-cause**<br>**(n=52)** | **unknown-cause**<br>**(n=34)** | **p-value** |
| --- | --- | --- | --- | --- |
| **Initial Rhythm at CPR**<br>**(81/86 patients), n(%)** |  |  |  | 0.155 |
| - VF | 67 (78) | 37 (71) | 30 (88) |  |
| - VT | 5 (6) | 3 (6) | 2 (6) |  |
| - Asystole | 5 (6) | 3 (6) | 2 (6) |  |
| - PEA | 4 (5) | 4 (8) | 0 (0) |  |
| **Repolarisation at baseline**<br>**ECG (79/86 patients), n(%)** |  |  |  | **0.020** |
| - ST-elevation | 13 (16) | 12 (26) | 1 (3) |  |
| - ST-depression | 4 (5) | 1 (2) | 3 (9) |  |
| - terminal negative T | 11 (14) | 4 (9) | 7 (22) |  |
| - preterm. negative T | 9 (11) | 7 (15) | 2 (6) |  |
| **Events in patient history,**<br>**n(%)** |  |  |  | 0.507 |
| - VT | 2 (2) | 2 (4) | 0 (0) |  |
| - Frequent ES | 3 (3) | 1 (2) | 2 (6) |  |
| - BBB | 2 (2) | 1 (2) | 1 (3) |  |
| **Events during hospitalisation, n(%)** |  |  |  | 0.646 |

|  |  |  |  |
| --- | --- | --- | --- |
| - VF | 5 (6) | 4 (8) | 1 (3) |
| - VT | 6 (7) | 3 (6) | 3 (9) |
| - Asystole | 1 (1) | 1 (2) | 0 (0) |
| - PEA | 0 (0) | 0 (0) | 0 (0) |
| - AF | 2 (2) | 1 (2) | 1 (3) |
| **Events after discharge, n(%)** |  |  |  | 0.444 |

|  |  |  |  |
| --- | --- | --- | --- |
| - VF | 5 (6) | 2 (4) | 3 (9) |
| - VT | 2 (2) | 1 (2) | 1 (3) |
| - AF | 1 (1) | 0 (0) | 1 (3) |

*AF: atrial fibrillation, BBB: bundle branch block, CPR: cardio-pulmonary resuscitation, ES:*
2 *extrasystole, PEA: pulseless electrical activity, VF: ventricular fibrillation, VT: ventricular*
3 *tachycardia.*

According to patient files, a total of 7 patients (8%) showed a history of arrhythmological conditions prior to the index event. During hospitalization, arrhythmic episodes were recorded in 14 patients (16%). In the aftermath of index hospitalization, over a median observation period of 3.3 years (IQR: 1.8-6.7), new episodes of arrhythmia were documented in 8 patients (9%). Between unknown-cause and definable-cause OHCA patients, no significant difference concerning peri-CPR rhythmological events was shown (before: p=0.494, during hospitalization: p=0.646, afterwards: p=0.444). The same accounts between patients with and without MAD (before: p=0.733, during hospitalization: p=0.912, afterwards: p=0.304).

### Laboratory Analysis

Within laboratory parameters, unknown-cause OHCA patients showed significantly lower values of serum creatinine (difference: 0.25mg/dl, p<0.001), calcium (0.1mmol/l, p=0.009), peak troponin T (409ng/ml, p=0.017) and peak N-terminal pro-brain-type natriuretic peptide (565ng/l, p=0.048). Results of lab analyses are shown in table 1.

## Discussion

This study is the first to investigate the role of MAD particularly in OHCA patients undergoing CMR imaging. Our results can be summarized as follows: a) MAD is a common feature in patients with unknown-cause OHCA, whilst b) it is far less common in patients with a definable cause of OHCA; c) MAD-patients in our study showed generally less comorbidities for cardiovascular events, however d) MAD was revealed to be an independent marker for unknown-cause OHCA after adjustment for age, hypertension and hypercholesterolemia.

### Prevalence of unknown-cause OHCA

In the present analysis, no definite cause for CA could be found in 40% of OHCA patients despite profound diagnostics. This number exceeds the observations of a German register study investigating 33,772 OHCA patients between 2007 and 2017. In that study, the proportion of unknown-cause OHCA was 17% (16). A possible explanation for this discrepancy is the fact that in most cases of definable-cause OHCA (e.g. myocardial infarction), there is no general recommendation for further investigation via CMR (17). Furthermore, due to its limited availability, CMR is usually only performed when the most common reasons for CA can be excluded beforehand. This additionally increases the percentage of unknown-cause OHCA in our study, in which, however, performance of CMR is a central inclusion criterion.

### MAD in unknown-cause OHCA

In this study, MAD was defined as end-systolic disjunction extent of at least 1mm, referring to an important forerunner-study by Dejgaard et al. (3). This approach can currently be regarded as a quite strict definition of MAD, as many other studies tended to define MAD as disjunction of any extent (6, 11, 18). However, in some rare studies, also larger cut-offs can be found, such as 2mm (19) and 5mm (20). As the minimum MAD in this present study was 2.8mm, shifting the threshold to 2mm would have had no effect on the outcome; however, a threshold of 5mm would have decreased the MAD-prevalence to 17% (n=15, 12 with unknown-cause OHCA, 35% vs. 3 with definable-cause OHCA, 6%, p<0.001).

One main finding of our study was that MAD was diagnosed significantly more often in unknown-cause OHCA patients, while these patients generally showed distinctly less comorbidities, especially in terms of age, BMI, blood pressure, hypercholesterolemia and CAD-prevalence. According to a cohort study by Essayagh et al. in 595 MVP-patients, the presence of disjunction was an independent risk factor for the occurrence of arrhythmic events in the long term (21). This finding is in line with a study by Dejgaard et al., which found severe arrhythmic events in 12% of MAD-patients and postulated MAD to be an arrhythmogenic risk factor itself, independent of concomitant prolapse (3). Accordingly, multivariate logistic regression analysis in the present study revealed MAD to be an independent marker of OHCA of unknown cause after adjustment for age, hypertension and hypercholesterolemia. There are hardly any other data available about the role of MAD in OHCA. However, a study by Lee et al. investigating the association of MVP and severe arrhythmias indicated that systolic curling motion in MAD was a strong and independent predictor of these events (15). In the present study, systolic curling motion was more common in unknown-cause OHCA patients; however, this difference was not significant, which is probably due to the small number of MAD in definable-cause OHCA patients.

### Features of unknown-cause OHCA

Besides MAD, female sex has proven to be a strong prognostic marker for unknown-cause OHCA, with 74% of these patients being female. Referring to the above-mentioned register study, almost 65% of all OHCA patients were male, which is in line with our study (58%). However, the percentage of women with unknown-cause OHCA in that register study was 40%. This is most likely due to the high rate of cardiac events in the definable-cause group (83%), which is accordingly more common in men (16). Then, although patients in the unknown-cause group in general had structurally normal hearts, some of them still showed an EF below 40%. A very sensible explanation for this phenomenon can be found in a study by Gonzalez et al., describing a marked decrease of LV-EF up to 25% due to cardiac arrest, hinting that perhaps these patients with a lower EF at CMR had a normal ventricular function pre-CPR (22). Additionally, it can be assumed that the partly quite short interval between CPR and CMR also plays a non-neglectable role here, as the LV function underlies a high variability during the first few days after cardiac recovery, which was shown by Kalra et al. in OHCA patients via echocardiography (23). Another finding, which is probably a result of the high frequency of cardiac triggers for CA in the present study is that LGE was significantly less common in unknown-cause OHCA and in MAD-patients. LGE was found to be a strong predictor for definable-cause OHCA. The percentage of patients with LGE in the definable-cause group (72%) is in line with a study by Neilan et al., detecting LGE in 71% of a patient cohort of 137 CA survivors (24). Contrary, the proportion of patients showing LGE in the unknown-cause group was less than a tenth. This could be due to the young age of patients in this group as well as the low risk profile.

### Epidemiologic features of MAD

In the present study, MAD was evident in 28% of patients in at least one segment of the posterior mitral leaflet. This is approximately in line with three studies reporting the prevalence of MAD via transthoracic echocardiography in MVP-patients (MAD in 22%) (25), via 3D-TEE in a mixed-patients cohort (27%) (15) and via CMR in myxomatous mitral valve disease (35%) (6). However, according to a recently published study by Toh et al., investigating the prevalence of MAD in a population of 98 patients without structural heart disease via CT, the true prevalence of MAD could be up to 96% (5). This marked difference to our present study could be at least partly due to the underlying examination method, as CT shows a higher spatial resolution than CMR, which also manifests itself in a larger median MAD-extent in our study (5.7mm vs. 3.0mm in Toh et al.).

The ratio of 75% women in MAD-patients is in line with a study by Perazzolo Marra et al., describing MAD as a constant feature of arrhythmogenic MVP (4). According to a large investigation of MVP prevalence in the course of the Framingham Study, MVP in general was shown to be a feature mainly affecting young women (26). As MAD is very often still accompanied by MVP, this would be in agreement with our data. However, data about sex distribution in MAD are currently rather inconsistent (2) and studies are still lacking.

### MAD and Arrhythmias

Interestingly, the difference in initial ECG findings at CPR between patients with and without MAD was of borderline significance, with all 24 MAD-patients showing VF at first medical contact. Overall, 84% of patients in the present study initially presented with a shockable rhythm, which is in line with a study by Majewski et al. investigating 871 OHCA patients that survived the first 30 days after CPR (27). In the first post-CPR-ECG, the main difference between the definable-cause and the unknown-cause group lies in the presence of repolarization disorders, being primarily due to the high ratio of myocardial infarctions showing ST-elevation in the first group, with the remaining entities being rather unspecific repolarization abnormalities. There are no data available about arrhythmias requiring CPR in MAD; however, as this study’s MAD-patients were at a distinctly younger age than patients without MAD and showed significantly less risk factors (i.e. diabetes, hypertension, hypercholesterolemia, CAD), these findings hint that the disjunction itself bears arrhythmogenic potential, especially in favour of ventricular arrhythmias (3). Further, although being more and more regarded as an arrhythmogenic entity itself, it is not yet clear why the presence of MAD seemingly predisposes for the development of severe arrhythmic events. Some studies postulate fibrosis of the papillary muscles as well as myocardial stretch by a contingently prolapsing leaflet as the primary pathophysiology of the MAD arrhythmic syndrome (28, 29). Another hypothesis involving damage or tissue change of the cardiac conduction system has not yet been sufficiently investigated (3, 30), but would on the other hand explain the increased risk of arrhythmic events in MAD-patients even without presence of MVP (3) or, as shown in our study, LGE. The fact that only 9% of unknown-cause patients in our study showed replacement fibrosis is probably due to a compound of these and maybe still unknown pathophysiological mechanisms that result in arrhythmias even before being measurable.

#### Limitations

We acknowledge that this study bears some limitations, with the most important being its retrospective nature, which results in partly incomplete patient history records and further course after discharge. Furthermore, the selected patient population is subject to a certain selection bias, as the percentage of patients with unknown-cause OHCA was overproportionally high, due to the availability of CMR imaging being a central inclusion criterion and the less common referral to CMR in definite-cause OHCA. Then, a probably very helpful tool and valuable addition to our analysis concerning tissue characterization in MAD could be parametric myocardial mapping. However, these sequences were not obtained in many patients due to these patients having been scanned before mapping sequences were commercially available. Lastly, CMR protocols were not entirely uniform due to the fact that CMR was primarily performed as a part of clinical routine rather than a scientific study; however, all patients were adequately evaluable in terms of MAD and cardiac function.

#### Conclusion

MAD is a common feature in OHCA patients without a definable substrate for CA. MAD-patients were younger, more often female and typically presented with a lower risk profile. However, the mere presence of MAD seems to be an independent factor of OHCA without clear trigger. Further research to characterize and understand the role of MAD in CA is needed.

## Data Availability

The data underlying this article will be shared on reasonable request to the corresponding author.

## List of Abbreviations

BMI: Body Mass Index
BSA: Body Surface Area
CA: Cardiac Arrest
CAD: Coronary Artery Disease
CI: Confidence Interval
CMR: Cardiovascular Magnetic Resonance Imaging
CPR: Cardio-Pulmonary Resuscitation
CT: Computed Tomography
CTA: Computed Tomography Angiography
CX: Circumflex Artery
ECG: Electrocardiogram
EDV: Enddiastolic Volume
EDVi: Enddiastolic Volume Indexed by Body Surface Area
ESV: Endsystolic Volume
ESVi: Endsystolic Volume Indexed by Body Surface Area
IQR: Interquartile Range
LAD: Left Anterior Descending Artery
LGE: Late Gadolinium Enhancement
LV: Left Ventricle/Ventricular
MAD: Mitral Annular Disjunction
MM: Myocardial Mass
MVP: Mitral Valve Prolapse
OHCA: Out-of-Hospital Cardiac Arrest
OR: Odds Ratio
RCA: Right Coronary Artery
VF: Ventricular Fibrillation

## Acknowledgements, Funding and Data Availability

This study did not receive any specific grant from funding agencies in the public, commercial, or not-for-profit sectors.

There is no conflict of interest.

The data underlying this article will be shared on reasonable request to the corresponding author.

